# History of SARS-CoV-2 infection, anti-spike IgG antibody kinetics and neutralization capacities following the second and third dose of BNT162b2 vaccine in nursing home residents

**DOI:** 10.1101/2022.02.07.22270557

**Authors:** H. Jeulin, C. Labat, K. Duarte, S. Toupance, G. Nadin, D. Craus, I. Georgiopoulos, I. Gantois, F. Goehringer, A. Benetos

**Author notes:** Address correspondence to: Prof. Athanase Benetos, MD, PhD, Head of the Department of Geriatrics, Hôpital Brabois, Rue Morvan, 54500, Vandoeuvre-lès-Nancy, University Hospital of Nancy (CHRU), Université de Lorraine, Nancy, France. The authors have declared no competing interest.

## Abstract

**Importance:** Duration of post-vaccination protection against COVID-19 in individuals is a critical issue, especially in nursing home (NH) residents, i.e. one of the most vulnerable populations.

**Objective:** To estimate the duration of the IgG(S) response to the mRNA BNT162b2 vaccine in NH residents with (COV-Yes) or without (COV-No) history of natural infection with SARS-CoV-2.

**Design, setting and participants:** IgG(S) quantification was carried out at 3 different time periods following administration of the Pfizer BioNtech vaccine: three then seven months after the 2^nd^ dose and one month after the 3^rd^ dose. 574 COV-Yes and COV-No NH residents were included in 2 cohorts: Main (n=115, mean age 84 years) or Confirmatory (n=459, mean age 88 years).

**Exposure:** All subjects received the BNT162b2 vaccine.

**Main outcomes and measurements:** IgG(S) antibodies and seroneutralization capacity.

**Results:** Neutralization capacity was strongly correlated with IgG(S) levels (R^2^:76%) without any difference between COV-Yes and COV-No groups for the same levels of IgG(S). COV-Yes, compared to the COV-No subjects showed 5-fold and 15-fold higher IgG(S) titers 3 and 7 months after the 2nd dose, but less than 2-fold higher IgG(S) after the 3^rd^ dose, due to a more pronounced effect of the 3^rd^ dose in the COV-No group. These results were similar in both cohorts. After the 2^nd^ dose, duration of assumed robust protection (IgG(S) >264 BAU/ml) was 2-fold higher in the COV-Yes vs. COV-No group: 12.60 (10.69-14.44) vs 5.76 (3.91-8.64) months, and this advantage was mainly due to the higher IgG(S) titers after the 2^nd^ dose and secondary to a slower decay over time. After the 3^rd^ dose, duration (months) of robust protection was estimated at 11.87 (9.88-14.87) (COV-Yes) and 8.95 (6.85-11.04) (COV-No).

**Conclusions and relevance:** In old subjects living in NH, history of SARS-CoV-2 infection provides a clear advantage in the magnitude and duration of high IgG(S) titers following the 2^nd^ dose. Importantly, the 3^rd^ dose induces a much more pronounced IgG(S) response than the 2^nd^ dose in COV-No subjects, the effect of which should be able to ensure in these subjects a prolonged protection against severe forms of COVID-19.

## Introduction

Mass vaccination of nursing home (NH) residents, the population most likely to develop severe forms of COVID-19, has resulted in an impressive decrease in SARS-CoV-2 contaminations as well as a dramatic decrease in COVID-19-related mortality (1). In France, NH residents received mRNA vaccines (mainly BNT162b2, Pfizer BioNtech) and the vast majority of them (81%) were fully vaccinated between January and June 2021 (2), while the 3^rd^ dose was administered between October and December 2021. Clinical studies in NHs demonstrated that, after the second dose, this very old population was able to develop SARS-CoV-2 IgG(S) antibodies, thus obtaining protection against severe forms of COVID-19 (3,4), and that previous infection by SARS-CoV-2 increased the immunogenicity of the vaccines (4). This latter result has also been observed in younger adults (5–7).

One of the most critical issues is the duration of post-vaccination protection in individuals with or without history of natural SARS-CoV-2 infection (7–9). The waning in serum SARS-CoV-2 antibodies has raised pressing questions regarding long-term immunity thus leading to the 3^rd^ dose vaccination.

In the present study, we investigated the IgG(S) response to the BNT162b2 vaccine in NH residents with or without history of SARS-CoV-2 infection at three different checkpoints: a) approximately three months after the leading 2 doses of the vaccine, b) four months later just before the booster (3^rd^) vaccine and c) one month after the booster vaccine. In addition, a sero-neutralization assay was performed for a subgroup of subjects and the results analyzed according their COVID-19 status and IgG(S) levels.

The results of these analyses should enable modelizing the decrease over time of IgG(S) SARS-CoV-2 antibodies and provide an estimate of the duration of protection of the 3^rd^ dose of the mRNA vaccine against severe forms of COVID-19 in this very old population.

## MATERIAL AND METHODS

### Participants

Two cohorts were formed (Main and Confirmatory) comprised of NH residents from the Nancy-Lorraine region with (COV-Yes) or without (COV-No) history of prior SARS-CoV-2 infection. Residents belonging in the Main cohort were included between April 17, 2021 and September 21, 2021 in 5 NHs, whereas subjects belonging in the Confirmatory cohort were included between August 10, 2021 and December 15, 2021 in 12 NHs (Supplemental Table 1). In the Main cohort, two IgG(S) quantifications were performed at mean 3 months (1^st^ IgG(S) quantification) and 7 months (2^nd^ IgG(S) quantification) following the 2^nd^ vaccination, whereas in the Confirmatory cohort, a single IgG(S) quantification was performed 7 months after the 2^nd^ vaccination (also defined as 2^nd^ IgG(S) quantification). In both cohorts, a 3^rd^ IgG(S) quantification was performed 1.5 months after the 3^rd^ (booster) dose (Figure 1). Age, sex, vaccination dates, SARS-CoV-2 infection dates, as well as IgG(S) quantification dates were recorded for all residents in both cohorts. In addition, for participants in the Main cohort, BMI, autonomy status and number of medications were also collected. This study was registered in ClinicalTrials.gov (NCT04964024) and received the approval of the Ethics Committee of the Nancy University Hospital (CHRU) (Comite d’Ethique CHRU de Nancy, decision n°326, August 3^rd^, 2021).

**Figure 1:**
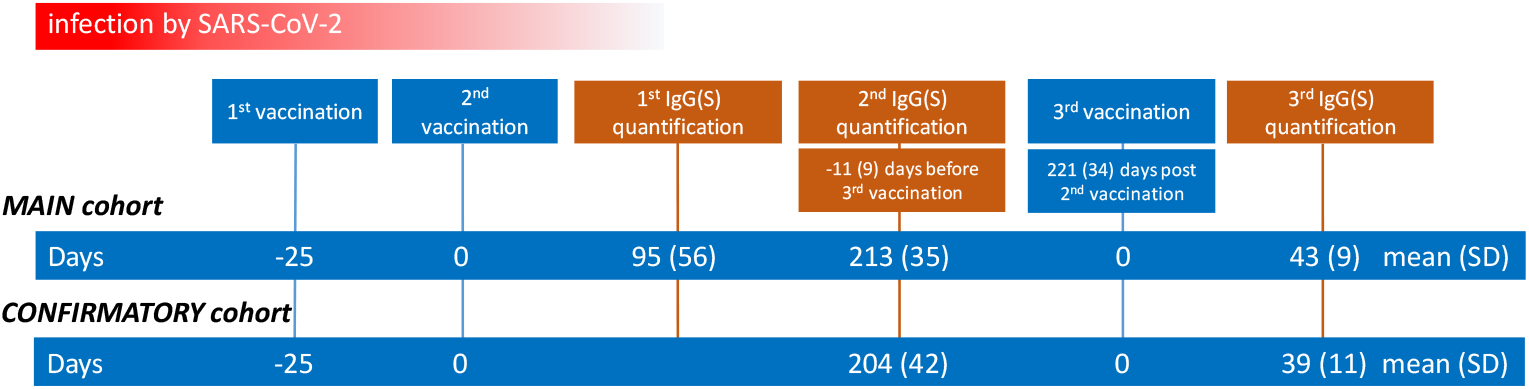
Number of days (D) between the 2^nd^ vaccination* and the 1st and 2^nd^ IgG(S) quantifications or the number of days between the 3^rd^ vaccination and the 3rd IgG(S) quantification in the Main and Confirmatory cohorts * For the 27 subjects who had SARS-CoV-2 infection a few days after the 2^nd^ vaccination (see legend figure 2), time to positive RT-PCR was taken into account in lieu of the 2^nd^ vaccination.

#### Inclusion criteria for both cohorts

i. NH residents aged 65 and older, with or without prior SARS-CoV-2 infection. Previous infection was identified either by history of positive RT-PCR or by anti-nucleoprotein (N) IgG quantification.
ii. SARS-CoV-2 anti-spike IgG(S) antibody quantification performed using the same quantification method (10) in the same laboratory.
iii. Complete vaccination scheme, meaning two injections of the BNT162b2 mRNA vaccine with the second injection at least 12 days before the first antibody quantification.
iv. Consent of the NH residents or their tutors for the use of the clinical and biological results for the aims of this study.

### Method for IgG(S) quantification

Blood samples were centrifuged to collect serum, the latter of which was stored at −20°C. Anti-spike IgGs were detected using the LIAISON® SARS-CoV-2 TrimericS IgG assay (Diasorin SA, France) on a Liaison XL Device (Diasorin SA, France), based on recombinant Trimeric Spike glycoprotein as capture antigen (10). Quantitative results are expressed as Binding Antibody Units (BAU/mL) according to the WHO first International Standard (IS) for anti-SARS-CoV-2 immunoglobulin (NIBSC code 20/136). Positive threshold was ≥ 33.8 BAU/mL. Samples with antibody titer >2080 were diluted to 1:20 as recommended by the manufacturer in order to determine accurate IgG levels. For technical reasons the samples of 14 patients of the Main and 6 patients from the Confirmatory cohort were not further diluted and these subjects were considered to have the plateau value of 2080 BAU/ml.

### Microneutralization assay

A sub-population of 39 NH residents was selected from the entire population according to IgG(S) levels following the 2^nd^ vaccination (“low”<650 BAU/ml vs. “high”>2080 BAU/ml) and prior SARS-CoV-2 infection (COV-Yes vs. COV-No) to assess serum neutralization capacities. Thus, 4 groups were studied: COV-No/low IgG(S), n=10, median 215 BAU/ml; COV-Yes/low IgG(S), n=10, median 217 BAU/ml; COV-No/high IgG, n=9, median 3270 BAU/ml; and COV-Yes/high IgG, n=10, median 4486 BAU/ml.

The SARS-CoV-2 B.1.617.2 (Delta) strain from a positive respiratory sample (Covi-Lor collection, Nancy University Hospital, France) was cultured on Vero E6 cells. Sera positive for anti-SARS-CoV-2 antibodies were diluted from 1/10 to 1/640 and incubated with live-virus suspension for 2 hr. Cells were inoculated with the final suspension. Each dilution was tested five times in each experiment and each sample was tested in two independent experiments. The cytopathic effect was read on day +6.

Negative controls consisted of uninfected cells while positive controls consisted of the virus incubated without sera and virus incubated with SARS-CoV-2–negative sera at a 1/10 ratio. The samples were classified according to neutralization activity at the 1:40 dilution, i.e. neutralization > 50% (NT50).

### Statistical analyses

Descriptive IgG data are presented as medians (IQR) and as mean (SD) values or percentages for the other variables. Due to the absence of normal distribution of IgG(S) values, comparisons between the different groups were performed after logarithmic transformation. Comparisons were conducted with ANOVA tests. Age and time (Δ) between the last immune stimulation (COVID-19 or vaccine) and antibody quantification were used for the adjusted models. Multiple regression analyses were also performed to test the role of clinical and demographic variables, time from immunization, and history of prior SARS-CoV-2 infection on IgG(S) levels. Pearson’s correlation was used to study the association between IgG(S) values and time of immunization for each studied group (see Figure 2). P <0.05 was considered statistically significant.

**Figure 2:**
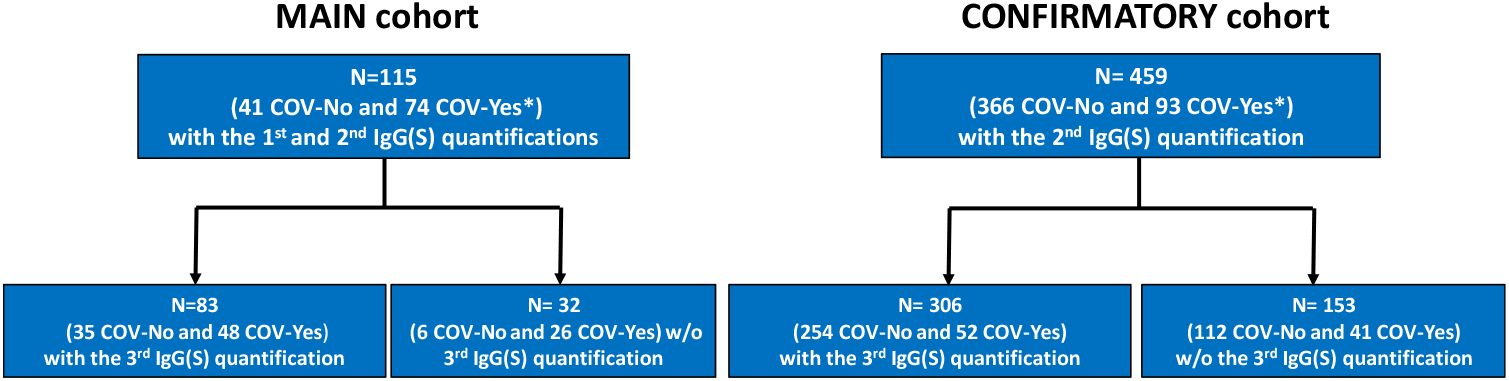
Study flowchart *Among COV-Yes subjects, 20 out of 74 in the Main cohort and 7 out of 93 in the Confirmatory cohort hadSARS-CoV-2 infection a few days after the 2^nd^ vaccine dose (February-March 2021). All other subjects had SARS-CoV-2 infection prior to the 1^st^ vaccine dose.

In an additional analysis NH residents were classified into 4 categories according to IgG(S) level in BAU/ml: ≥34 (level of positivity), ≥264 (threshold associated with higher protection against severe COVID-19 forms (11), ≥2080 threshold highly predictive of strong neutralization response (12) Differences between COV-No vs. COV-Yes were compared using a χ^2^ test.

#### Models used for the assessment of IgG decay with time

In both cohorts, the natural logarithm of the IgG rate follows a linear regression with steepness B(1), meaning that the decay rate of the IgG is exponential. Hence, the proportion q_30_ of IgG that is lost during one month is q_30_ = 1-exp(30*B(1)).

If the rate of IgG(S) decays exponentially by a proportion q_30_ each month, then if a quantity I(0) is measured at time t=0, the immunity will be lost at a time t_loss_ (in months) at which the IgG(S) rate equals I_loss_ given by t_loss_=−log(I(0)/I_loss_)/log(1-q_30_).

A linear mixed model with random effect on subjects was used to examine IgG kinetics after the second dose of vaccine in the Main cohort. The dependent variable consisted of the IgG(S) level, which was log-transformed. Fixed effect covariates included time after the second dose of vaccine in months as linear, SARS-CoV-2 status, and interaction between time and SARS-CoV-2 status. Given the low power of interaction tests (13,14), a significance level of 0.10 was used for interaction p-values. Similar analysis was performed in the Confirmatory cohort using a classical linear regression model.

## RESULTS

### Cohort characteristics

The Main cohort was comprised of 115 subjects (41 COV-No and 75 COV-Yes) whereas the Confirmatory cohort was comprised of 459 subjects (366 COV-NO and 93 COV-Yes) (Figure 2). Demographic characteristics and IgG(S) levels are presented in Table 1. BMI, autonomy status and number of daily medications collected in the Main cohort were similar in the COV-Yes and COV-No groups.

**Table 1:**
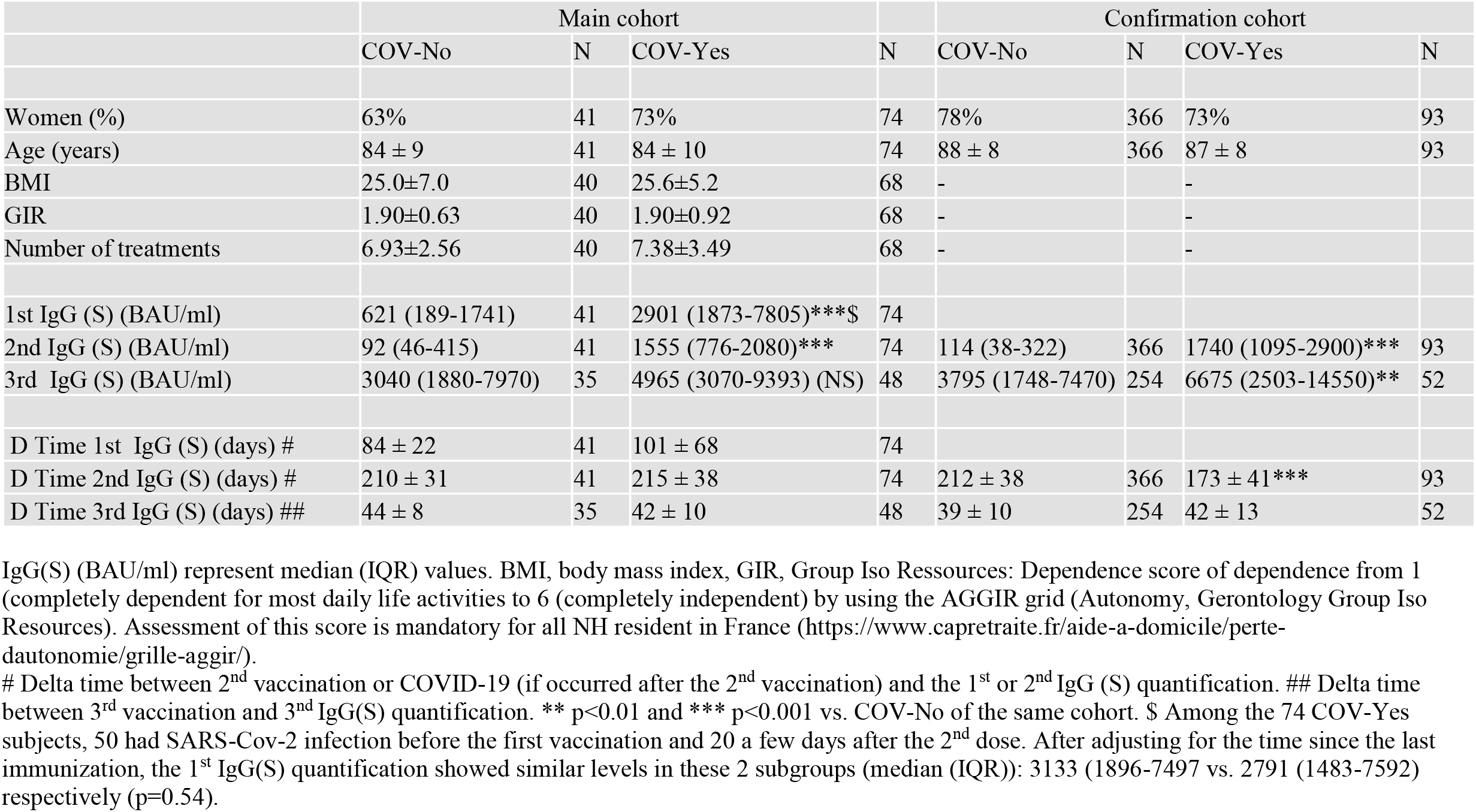
Clinical data and IgG(S) levels in the Main and Confirmatory cohorts in the two subgroups of NH residents with (COV-Yes) or without (COV-No) history of prior SARS-CoV-2 infection.

#### SARS-CoV-2 IgG(S) antibody quantification: comparison between COV-No vs. COV-Yes groups

In the Main cohort (Table 1, left), significantly lower IgG(S) titers were found in the COV-No vs. COV-Yes subjects for 1^st^ IgG(S) quantification (median: 621 vs. 2901 BAU/ml)) (p<0.001). No difference in IgG(S) (time-adjusted) levels were observed between the subjects who presented SARS-CoV-2 infection before or after the 2^nd^ vaccination (Table 1 legend). The same results were observed for the 2^nd^ IgG quantification for both cohorts (Table 1, right). After the booster dose (3^rd^ IgG quantification), IgG(S) increased dramatically in all groups. A significant difference in favor of COV-Yes was found only in the Confirmatory cohort (3795 vs. 6675 BAU/ml) (p<0.01). No differences in IgG(S) levels were observed between the Main and Confirmatory cohorts in each COVID-19 status group both for the 2^nd^ and 3^rd^ IgG quantifications. The COV-Yes/COV-No IgG(S) ratio after the 3^rd^ dose dramatically decreased compared to the ratio after the 2^nd^ dose (2^nd^ quantification) from 16.9 to 1.6 (Main cohort) and from 15.3 to 1.8 (Confirmatory cohort).

The evolution of the individual IgG(S) levels in the two groups of the Main cohort in residents having all 3 IgG(S) measurements showed that, in the COV-No group, the response to the 3^rd^ vaccination was significantly higher than the response to the 2^nd^ vaccination, whereas no such difference was observed in the COV-Yes group (Supplemental Figure 2). Figure 3 shows the classification of residents according to IgG(S) levels during the different quantifications: 95.1% of the COV-No and 100% of the COV-Yes residents of the Main cohort (upper panel) showed a positive SARS-CoV-2 (>34 BAU/ml) serology during the first IgG(S) quantification. A more pronounced IgG(S) response in the COV-Yes was clearly observed during the first and the second quantifications (p<0.0001) but not in the 3^rd^ especially in the Confirmatory cohort (p=0.47). Noteworthy, in the 2^nd^ quantification performed 7 months after the second dose, the percentage of residents with levels >264 BAU/ml, reached up to 93% in the COV-Yes group but only 28% in the COV-No groups of the Main and 92.5% and 28,2% in the Confirmatory cohort respectively.

**Figure 3:**
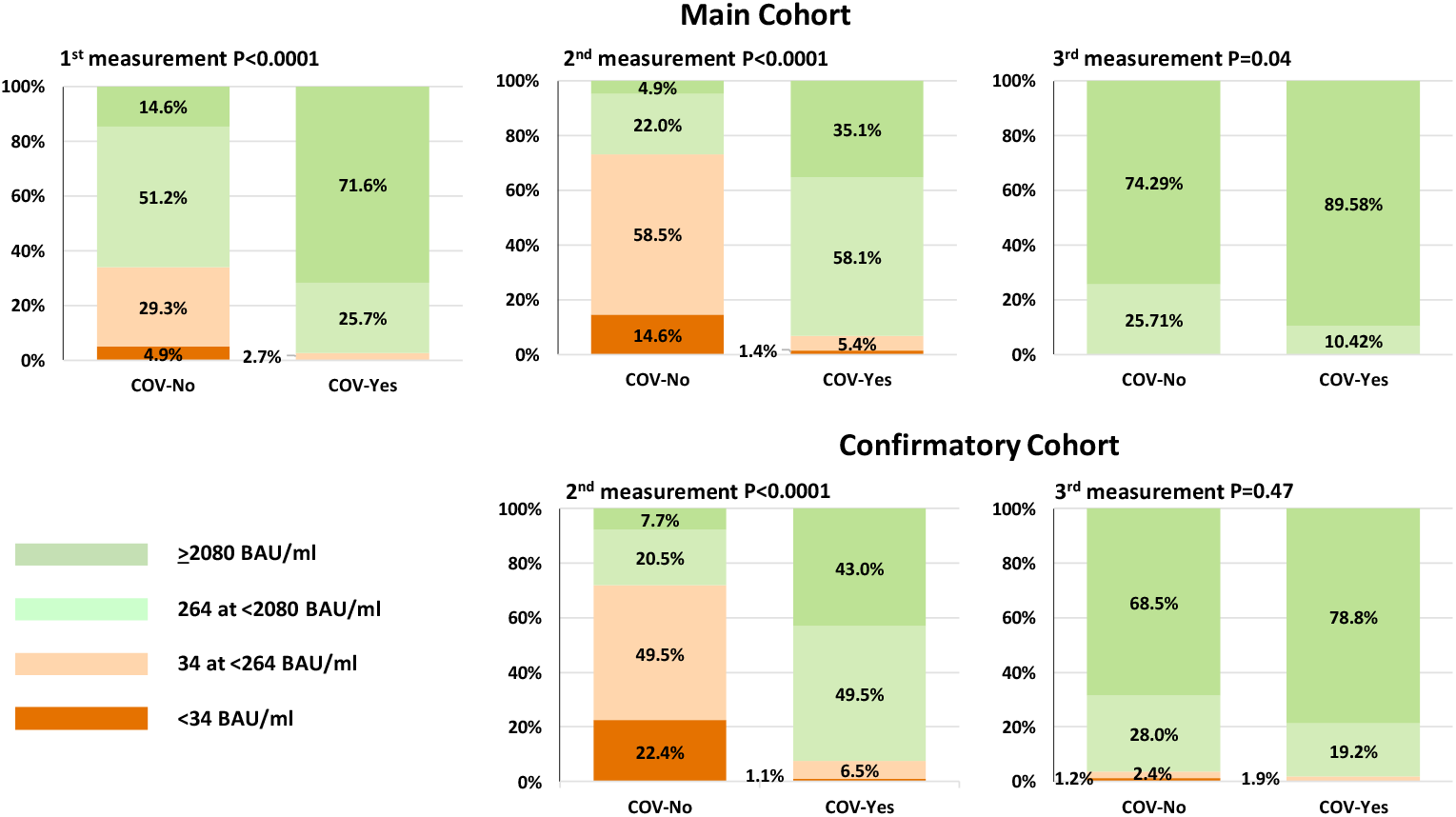
Classification of the subjects in 4 IgG(S) level categories (BAU/ml) during the 1^st^ (left), 2^nd^ (middle) and 3^rd^ (right) quantification according to history of SARS-CoV-2 contamination in the Main (upper) and the Confirmatory (lower) cohorts.

Multivariate analyses in the Main cohort revealed that IgG(S) levels during the first and the second quantifications were dependent on both SarS-CoV-2 status (p<0.00001) and time since last immunization (p<0.001). Age, sex and clinical parameters did not influence IgG(S) levels,.

### Microneutralization assay

A strong positive relationship was found between IgG(S) levels and neutralization activity (NT50) (R2=0.77, p<0.001) (Supplemental Figure 1). No difference was observed when comparing neutralization activity of post-vaccination serum in residents with or without history of prior COVID-19 (COV-Yes vs. COV-No: p=0.102 for low IgG(S) titers and p=0.567 for high IgG titers. Multivariate analysis, showed that neutralization activity was determined by IgG(S) levels (p<0.0001) but not by SARS-CoV-2 status (p=0.98).

Serum from NH residents with low-level IgG titers (< 264 BAU/mL) neutralized the virus *in vitro* (NT50 ≥40) in 2/17 (11.8%) cases, whereas serum from NH residents with high-level IgG titers (≥264 BAU/mL) neutralized the virus *in vitro* (NT50 ≥40) in all 22/22 (100%) cases (p<0.0001 vs. low IgG titers).

#### IgG waning rates in the Main and the Confirmatory cohorts

IgG decay over time followed an exponential pattern, which allowed establishing a linear model between the log of IgG and the time since the 2^nd^ vaccination (Table 2). In the Main cohort, the beta coefficients of the IgG/time relationship were highly significant for both groups (p<0.001) and the interaction between time and SARS-CoV-2 status group was significant (p=0.051). The proportion of IgG(S) lost during one month, estimated from the linear mixed model, was 29.5% (95% CI: 26.4-32.5) in the COV-No group and 25.4% (95% CI: 22.6-28.0) in the COV-Yes group. Figure 4 shows the distribution of IgG(S) levels according to time after the second dose and SARS-CoV-2 status in the Main and the Confirmatory cohorts.

**Tableau 2:** Linear regression models for the association with Log-IgG(S) after the second dose of BNT162b2 vaccine.

|  | Main cohort<br>(linear mixed effect model) |  | Confirmatory cohort<br>(linear model) |  |
| --- | --- | --- | --- | --- |
|  | Beta (SE) | p-value | Beta (SE) | p-value |
| History of SARS-Cov-2 infection |  |  |  |  |
| COV-No | 7.31 (0.21) | <0.0001 | 8.30 (0.47) | <0.0001 |
| COV-Yes | 9.21 (0.17) | <0.0001 | 9.20 (0.72) | <0.0001 |
| Time since second dose of vaccine (months) |  |  |  |  |
| Among COV-No subjects | -0.35 (0.02) | <0.0001 | -0.49 (0.07) | <0.0001 |
| Among COV-Yes subjects | -0.29 (0.02) | <0.0001 | -0.31 (0.12) | 0.011 |
| Time since second dose of vaccine x Covid-19 status | Interaction p-value = 0.051 |  | Interaction p-value = 0.20 |  |

**Figure 4:**
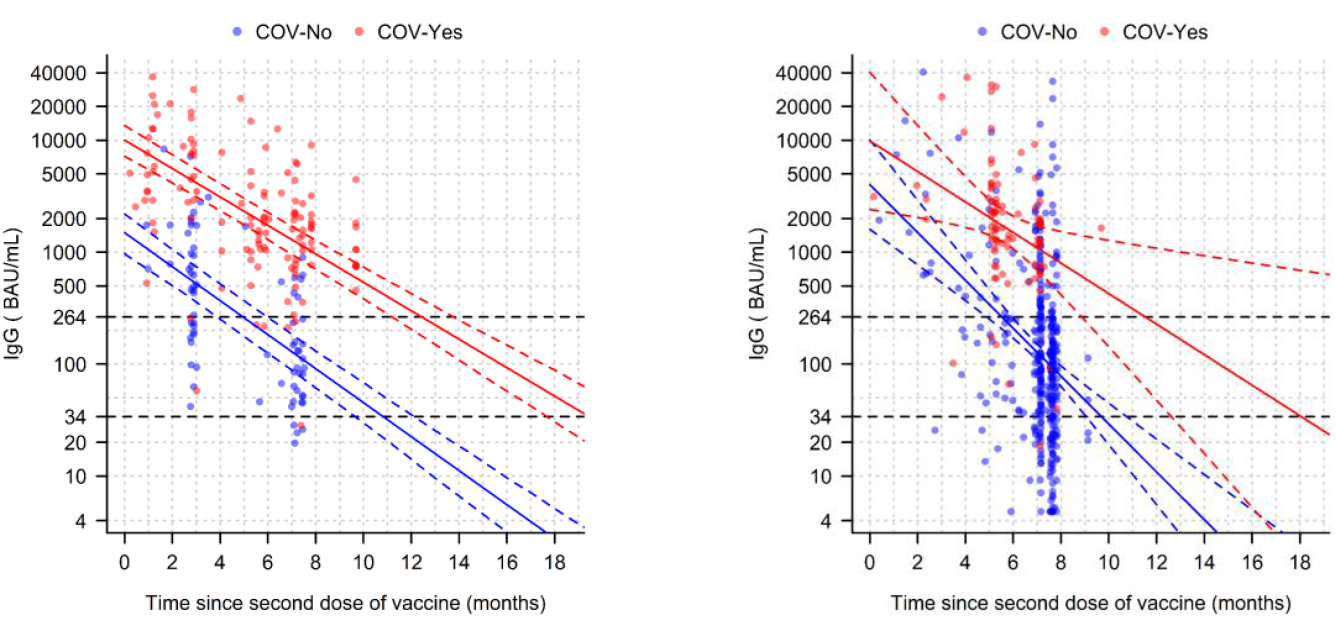
Distribution of IgG(S) levels according to time after the second dose of vaccine and COVID-19 status in the Main (left) and Confirmatory (right) cohorts IgG is represented in logarithmic scale. Solid lines represent the predictions from the model and dotted lines the 95% confidence interval. The horizontal black dotted lines correspond to the IgG(S) titers of negative serology (34 BAU/ml) and of “robust” protection (264 BAU/ml) In the Main cohort, each individual is represented twice corresponding to the 1^st^ and the 2^nd^ IgG(S) quantifications in accordance with the linear mixed model described in the Methods section

#### Estimation of immunization duration and robust protection following the 3^rd^ dose

For this estimation, the results of the IgG(S) decay in the Main (longitudinal) cohort were used with the assumption that the waning rates of IgG(S) over time in each COVID-19 status group were similar after the 3^rd^ dose as compared to the observed decay after the 2^nd^ dose. Following this analysis in the totality of the subjects who had received the 3^rd^ dose (n=389), the time (months) to return, under the threshold of 264 BAU/ml in the COVID-No group was estimated at 8.95 (6.85–11.04) (median (IQR)) after the 3rd dose vs. 5.76 (3.91–8.64) (p<0.00001) after the 2^nd^ dose and 11.87 (9.88–14.87) vs. 12.60 (10.69–14.44) (NS) in the COV-Yes group, respectively.

The corresponding times to return under the positivity levels (<35 BAU/ml) in COVID-No residents were estimated at 14.73 (12.63–16.83) vs. 11.54 (9.69–14.42) months (p<0.00001) and 18.77 (16.78–21.76) vs. 19.50 (17.58 – 21.34) months (NS) in the COVID-Yes residents, respectively.

For both the 2^nd^ and 3rd doses, these times were longer in the COV-Yes than the COV-No group (p<0.0001), while the difference in absolute values related to COVID status were also attenuated after the 3rd dose.

## Discussion

In the present study, we analyzed the IgG(S) response to the 2^nd^ and 3^rd^ doses of the Pfizer BioNtech mRNA vaccine in 2 different cohorts (Main and Confirmatory) comprised of a total of 574 NH residents. This design enabled us to show that the totality of the results observed in the Main Cohort were also observed in the Confirmatory cohort which, in our opinion, provides robustness to the data and conclusions presented herein.

The study’s focus was aimed at assessing the IgG(S) response after the 2^nd^ and the 3^rd^ vaccination since there is currently sufficient evidence regarding the importance of IgG(S) titers for the robust protection against severe forms of the disease, which is a major concern in this very old and highly frail population. In addition, in a subgroup of this population, we analyzed the relationship between IgG(S) and neutralizing capacity in residents with or without prior SARS-CoV-2 infection.

Two methodological aspects of the present study are particularly noteworthy:

-The method used for antibody quantification involved a whole recombinant trimeric protein for the detection of IgG (10). This method therefore allows detecting all antibodies directed against the S protein, and not only those linked to the RBD, the ACE2 binding site. The quantification range of this method is linked to the WHO standard, thus allow determining the IgG(S) titer in BAU/mL (15).
-Similarly, the conventional microneutralization method used herein was based on the original live SARS-CoV-2 virus to better approximate *in vivo* humoral immunity. Due to their characteristics, the results of these two methods are expected to be correlated and allow a relevant approach to the correlate of protection of anti-SARS-CoV-2 IgG(S). Thus, as shown in previous reports (3,8) the neutralization capacity was strongly correlated with IgG(S) levels in both COV-Yes and COV-No subjects. Of particular interest is that for similar levels of IgG(S), no difference between COV-No vs. COV-Yes was observed.

A number of clinical studies conducted in NH have shown that residents with a history of SARS-Cov-2 infection showed a more pronounced post-vaccination IgG(S) level than residents without prior infection (3,4). Our study confirms this significant difference and further adds key information showing that 7 months after the 2^nd^ dose, this difference was even more pronounced, i.e. the COV-Yes group showed median IgG(S) titers 15-fold higher than the COV-No group, in both the Main and Confirmatory cohorts. In addition, the percentage of subjects with IgG(S) titers >264 BAU/ml, was only 28% in COV-No residents of the both cohorts, whereas among COV-Yes residents, approximately 93% in both cohorts featured IgG(S) titers >264 BAU/ml which seems associated to robust protection (11). These results indicate that among NH residents, the risk of loss of immunity over a relatively short period following the initial 2-dose vaccination is much higher in subjects without prior SARS-CoV-2 infection.

As also shown previously (16), our analyses revealed that in both COVID status groups, the decay in IgG(S) followed an exponential pattern, which allowed obtaining a linear relationship between the log-IgG(S) and time interval after the second vaccination. These analyses showed that COV-Yes residents presented a duration of immunization more than twice that of COV-No residents, primarily due to the much higher IgG(S) levels after the second vaccination in the COV-Yes group and secondarily due to the lower rates of IgG(S) waning in COV-Yes individuals.

Of note, following the 3^rd^ vaccination, the IgG(S) levels were higher than those observed after the 2^nd^ dose in COV-No residents, thereby leading to an approximate 50% increase in the duration of robust protection in this group after the 3^rd^ vaccination, although this effect was still less pronounced than the duration of robust protection of the 3^rd^ dose in the COV-Yes group. These results suggest that in NH residents without prior history of SARS-CoV-2 infection, protection against the severe forms of COVID-19 with the 3rd dose will be considerably longer compared to the 2^nd^ dose and similar to the protection obtained in COV-Yes subjects after the 2nd vaccination.

This study has two main limitations: first, the definition of COV-Yes by PCR and/or IgG(N)+ cannot eliminate that some subjects considered as being COV-No, had indeed a history of previous asymptomatic SARS-Cov-2 infection (7). Second, only exploration of humoral but not cellular immunity was performed, which probably has an important role in the prevention of serious disease forms. Moreover, the emergence of omicron or other variants in the future may modify the duration of protection.

In conclusion, in this very old population of NH residents, previous SARS-Cov-2 infection induces a more pronounced IgG(S) response to the BNT162b2 vaccine leading to a longer protection of the 2nd dose and in a lesser degree of the 3^rd^ dose. We anticipate than in NH residents without history of SARS-CoV-2, the 3^rd^ dose of the RNA vaccine, compared to the 2^nd^ dose, will induce a more prolonged protection against severe forms of COVID-19.

## Data Availability

All data produced in the present study are available upon reasonable request to the authors

## Acknowledgments

Tha authors thank the following medical coordinators of the participating NHs :Marc Berr, Thierry Collin, Caroline Ferry-Bert, Marie-Chrisine Godard, Patrick Lucquin, Lydie Osnowycz, Henri Rozenfarb; We thank Ms Anne Fréminet, Alice Metz and Cecile Lacomy for their valuable contribution. We thank all the directors and the staff of the 14 nursing homes for contributing to the realization of this study. We thank Mr Pierre Pothier for language review and stimulating discussions.

## References

1. Teran RA, Walblay KA, Shane EL, Xydis S, Gretsch S, Gagner A et al. Postvaccination SARS-CoV-2 infections among skilled nursing facility residents and staff members - Chicago, Illinois, December 2020–March 2021. MMWR Morb Mortal Wkly Rep. 2021;30;70:632–638.https://www.santepubliquefrance.fr/content/download/354025/3069653

2. Helle F, Moyet J, Demey B, François C, Duverlie G, Castelain S et al. Humoral anti-SARS-CoV-2 immune response after two doses of Comirnaty vaccine in nursing home residents by previous infection status. Vaccine. 2022;40:531–535.

3. Salmerón Ríos S, Cortés Zamora EB, Avendaño Céspedes A, Romero Rizos L, Sánchez-Jurado PM, Sánchez-Nievas G et al. Immunogenicity of the BNT162b2 vaccine in frail or disabled nursing home residents: COVID-A study. J Am Geriatr Soc. 2021;69:1441–1447.

4. Gaebler C, Wang Z, Lorenzi JCC, Muecksch F, Finkin S, Tokuyama M et al. Evolution of antibody immunity to SARS-CoV-2. Nature. 2021;591:639–644.

5. Stamatatos L, Czartoski J, Wan YH, Homad LJ, Rubin V, Glantz H et al. mRNA vaccination boosts cross-variant neutralizing antibodies elicited by SARS-CoV-2 infection. Science 2021;372:1413–1418.

6. Gallais F, Gantner P, Bruel T, Velay A, Planas D, Wendling MJ et al. Evolution of antibody responses up to 13 months after SARS-CoV-2 infection and risk of reinfection. EBioMedicine 2021 Sep;71:103561. doi: 10.1016/j.ebiom.2021.103561.

7. Levin EG, Lustig Y, Cohen C, Fluss R, Indenbaum V, Amit S et al. Waning Immune Humoral Responseto BNT162b2 Covid-19 Vaccine over six Months. N Engl J Med. 2021; 385:e84. DOI: 10.1056/NEJMoa2114583.

8. Zhong D, Xiao S, Debes AK, Egbert ER, Caturegli P, Colantuoni E et al. Durability of Antibody Levels After Vaccination With mRNA SARS-CoV-2 Vaccine in Individuals With or Without Prior Infection. JAMA 2021;326:2524–2526.

9. Bonelli F, Blocki FA, Bunnell T, Chu E, De La O A, Grenache DG al. Evaluation of the automated LIAISON® SARS-CoV-2 TrimericS IgG assay for the detection of circulating antibodies. Clin Chem Lab Med 2021;59:1463–1467.

10. Feng S, Phillips DJ, White T, Sayal H, Aley PK, Bibi S et al. Correlates of protection against symptomatic and asymptomatic SARS-CoV-2 infection. Nat Med. 2021;27:2032–2040.

11. Meschi S, Matusali G, Colavita F, Lapa D, Bordi L, Puro Vet al. Predicting the protective humoral response to a SARS-CoV-2 mRNA vaccine. Clin Chem Lab Med. 2021;59:2010– 2018.

12. Greenland S. Tests for interaction in epidemiologic studies: a review and a study of power. Stat Med. 1983;2:243–251.

13. Brookes ST, Whitely E, Egger M, Smith GD, Mulheran PA, Peters TJ. Sub-group analyses in randomized trials: risks of subgroup-specific analyses; power and sample size for the interaction test. J Clin Epidemiol. 2004;57:229–236.

14. Scheiblauer H, Nübling CM, Wolf T, Khodamoradi Y, Bellinghausen C, Sonntagbauer M. et al. Antibody response to SARS-CoV-2 for more than one year kinetics and persistence of detection are predominantly determined by avidity progression and test design. J Clin Virology. 2022;146:105052.

15. Khoury DS, Cromer D, Reynaldi A, Schlub TE, Wheatley AK et al. Neutralizing antibody levels are highly predictive of immune protection from symptomatic SARS-CoV-2 infection. Nat Med. 2021;27:1205–1211.

