## supplemental figure 1 and 2 and table 1 for "History of SARS-CoV-2 infection, anti-spike IgG antibody kinetics and neutralization capacities following the second and third dose of BNT162b2 vaccine in nursing home residents"

Seroneutralization activity (NT50) according to IgG(S) levels in sera from subjects with (COV-Yes) or without (COV-No) history of prior SARS-CoV-2 infection (upper panel) and multivariate regression analysis to explain variations in seroneutralization activity according to IgG(S) levels (post-second vaccination) and presence of history of SARS-CoV-2 (lower panel).

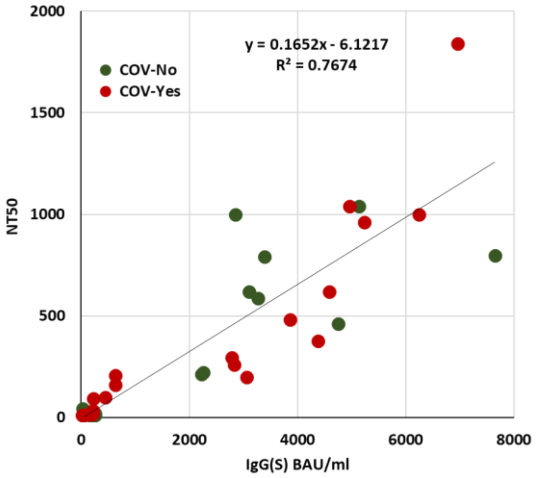

**Multivariate regression**

**Dependent Variable: NT50**

| | **Beta (SE)** | **$R^2$** | **P** |
|---|---|---|---|
| **IgG (1 BAU/ml)** | 0.1765 (0.015) | 75.6% | <0.0001 |
| **COVID (yes)** | -1.404 (68.758) | 0.0% | 0.98 |
| **Model** | | 76.7% | <0.0001 |

The strong positive relationship found between IgG(S) levels and neutralization activity (NT50) using a Pearson bivariate analysis was confirmed in the multivariate regression analysis. This multivariate analysis showed no influence of the SARS-CoV-2 status on the seroneutralization activity.

**Supplemental Figure 2**:

Evolution of the individual IgG(S) levels in the Main cohort in COV-Yes (red, n=35) and COV-No (green, n=34) residents having all 3 IgG(S) quantifications. Median values are in bold lines.

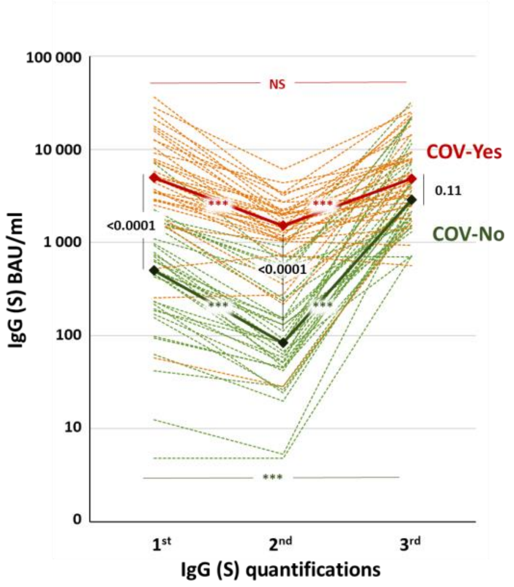

***Comparison between 2nd and 3rd vaccinations in the COV-No and COV-Yes NH residents.*** These data show that in the COV-No group, the response to the 3rd vaccination was significantly higher than the response to the 2nd vaccination, whereas no such difference was observed in the COV-Yes group. Since the time delay between the last immunization and IgG quantification differed in the 1st (mean 95 days) and 3rd IgG quantifications (mean 43 days) (see Figure 1), these values were adjusted for this time delay taking into account the IgG waning model (exponential) described above. This analysis confirmed a significant amplification in the IgG(S) response following the 3rd vaccination in the COV-No subjects (p<0.001) but no difference between the 2nd and 3rd vaccinations in the COV-Yes individuals (p=0.55). Subjects having IgG(S) values >2080 BAU/ml and for whom further dilutions could not be performed to obtain an exact IgG quantification (n=14) were excluded from this analysis (no possibility to calculate changes in absolute values between the 3 IgG(S) quantifications). *** P<0.0001 (t-paired test between the different quantification in each group) and T test for comparison between COV-No vs COV-Yes in the main cohort.

**Supplemental Table 1:**

Number of residents per Nursing home, in the Main and the Confirmatory cohorts

| | **Cohort** | | **Total** |
|---|---|---|---|
| **Nursing Homes** | **Main** | **Confirmatory** | |
| Benichou Nancy | | 44 | 44 |
| Le Clos Pré Saint Max | | 20 | 20 |
| Einville au Jard | 4 | 14 | 18 |
| Hôtel Club Saint Max | | 74 | 74 |
| Joudreville les Bruyères | | 40 | 40 |
| L’Oseraie Laxou | | 30 | 30 |
| Pompey Lay Saint Christophe | 44 | 62 | 106 |
| Résidence le Parc | 5 | 44 | 49 |
| Pont-à-Mousson | | 43 | 43 |
| Les Sablons Pulnoy | 18 | 46 | 64 |
| Sainte Sophie Thiaucourt | | 22 | 22 |
| USLD CHRU Nancy | 44 | 20 | 64 |
| **Total** | 115 | 459 | 574 |
